# Chronic Pain in Parkinson’s Disease: Prevalence, Sex Differences, Regional Anatomy and Comorbidities

**DOI:** 10.1101/2025.02.09.25321969

**Authors:** Natalia S. Ogonowski, Freddy Chafota, Fangyuan Cao, Amanda Wei Yin Lim, Victor Flores-Ocampo, Santiago Díaz-Torres, Zuriel Ceja, Luis M. García-Marín, Scott F. Farrell, Kishore R. Kumar, Jane Alty, George D. Mellick, Trung Thành Ngô, Miguel E. Renteria

**Affiliations:** Brain & Mental Health Program, QIMR Berghofer Medical Research Institute, Brisbane, QLD, Australia; School of Biomedical Sciences, Faculty of Medicine, The University of Queensland, Brisbane, QLD, Australia; Population Health Program, QIMR Berghofer Medical Research Institute, Brisbane, QLD, Australia; RECOVER Injury Research Centre; NHMRC Centre of Research Excellence: Better Health Outcomes for Compensable Injury; STARS Education and Research Alliance; The University of Queensland and Surgical, Treatment & Rehabilitation Service (STARS), Metro North Health, Herston, Brisbane QLD 4029, Australia; Department of Neurology and Molecular Medicine Laboratory, Concord Repatriation General Hospital, Concord, NSW 2139, Australia; Sydney Medical School, University of Sydney, Camperdown, NSW 2050, Australia; Translational Neurogenomics Group, Genomics and Inherited Disease Program, Garvan Institute of Medical Research, Darlinghurst, NSW 2010, Australia; School of Clinical Medicine, UNSW Medicine & Health, St Vincent’s Healthcare Clinical Campus, Faculty of Medicine and Health, UNSW Sydney; Wicking Dementia Research and Education Centre, University of Tasmania, Liverpool Street, Hobart, TAS 7001, Australia; School of Medicine, University of Tasmania, Hobart, TAS 7001, Australia; Royal Hobart Hospital, Hobart, TAS 7001, Australia; Griffith Institute for Biomedicine and Glycomics, Griffith University, QLD 4111, Australia; School of Biomedical Sciences, Faculty of Health, Queensland University of Technology, Brisbane, QLD, Australia

## Abstract

**Objective:** Chronic pain is prevalent among people living with Parkinson’s disease (PD). We analysed data from 10,631 Australian individuals with PD to assess the prevalence, age and sex differences, severity, anatomical distribution, clinical history, and associated factors.

**Methods:** We analysed data from 10,631 participants with PD enrolled in the Australian Parkinson’s Genetics Study (APGS), an ongoing nationwide cohort. Participants completed an online or paper-based questionnaire assessing sociodemographic factors, PD-related variables, and chronic pain characteristics. Chronic pain was defined as pain persisting for >3 months and occurring most days or daily. Statistical analyses included descriptive statistics, correlation analyses, and group comparisons using Chi-squared tests, Fisher’s exact tests, and independent samples t-tests.

**Results:** Two-thirds (66.2%) reported chronic pain, with females experiencing higher prevalence (70.8%) and severity (4.7 vs. 4.3 on a 10-point scale). Common pain sites included the buttocks (35.6%), lower back (25.4%), neck (19.4%), and knees (17.2%). Chronic pain was strongly linked to comorbid depression, sleep disorders, and osteoarthritis (p < 0.05). Environmental exposures such as pesticides, heavy metals, and alcohol were associated with higher pain prevalence, especially in males (p < 0.05).

**Interpretation:** These findings emphasise the substantial burden of chronic pain in PD, highlighting sex differences and strong links to multimorbidity. Further research is warranted to clarify sex-specific treatments and identify novel therapeutic targets.

---

Parkinson’s disease (PD) is a progressive neurodegenerative disorder characterised by a range of motor and non-motor symptoms. In recent decades, the significance of non-motor symptoms in PD has increasingly drawn the attention of both clinicians and researchers^1,2^. In particular, chronic pain is one of the most prevalent, affecting between 30%–85% of individuals with PD^3,4^. Despite its high prevalence and substantial impact on patients’ quality of life, chronic pain in PD remains underdiagnosed and poorly managed in clinical settings^5^.

The pathophysiology of chronic pain in PD remains poorly understood but is thought to involve both peripheral and central mechanisms. Pain can emerge at any stage of the disease and may even precede the onset of motor symptoms, highlighting its significance in PD progression. Despite its prevalence, chronic pain in PD remains one of the least understood aspects of the disease, largely due to its heterogeneous presentation and the inherent challenge in measuring subjective experiences of pain^5^. Pain in PD encompasses several types, including musculoskeletal, neuropathic, dystonic, central nociceptive, neuropathic, and nociplastic pain classifications^6^. These types also often overlap, further complicating diagnosis and treatment^7^.

Specific demographic and clinical factors have been associated with the variability of chronic pain in PD. Age at onset^8^, sex^9^, ethnicity, and the presence of comorbid conditions can all shape the perception and experience of pain^9,10^. For example, females with PD may experience more severe or more frequent pain than males^9^. At the same time, people with younger-onset PD may report different pain patterns compared to those diagnosed later in life^11^. These observations highlight a critical gap in understanding how these factors interact to influence chronic pain in PD, thus emphasising the need for further investigation.

Here, we analysed data from 10,631 participants from a nationwide cohort of Australian adults living with PD to (i) determine the prevalence of chronic pain, (ii) examine age and sex differences associated with chronic pain, (iii) assess the severity and anatomical distribution of chronic pain among individuals with PD, and (iv) identify social and lifestyle factors linked to chronic pain variability. Together, these objectives aim to elucidate the relationship between chronic pain and PD as the foundation for further targeted studies into optimal diagnostic and therapeutic intervention approaches.

## METHODS

### Study design and participants

The Australian Parkinson’s Genetics Study (APGS) is an ongoing nationwide cohort study that aims to characterise the clinical, genetic, and phenotypic diversity of PD in Australia. The target recruitment is 20,000 participants, including 10,000 diagnosed with PD and 10,000 neurologically healthy controls without a positive family history of PD.

Recruitment was undertaken through a combination of assisted mailouts sent to Australian residents who have been prescribed medications with an indication for PD in the last five calendar years and a public outreach campaign leveraging traditional and social media channels. Participation entails completing an online questionnaire, which includes sections on living with PD, clinical history (with a specific focus on chronic pain), family history of neurological conditions, lifestyle, and environmental risk factors. The online survey is hosted on the Qualtrics platform, but participants without computer access or those preferring a paper format can request a traditional paper version. The recruitment strategy of APGS has been detailed elsewhere ^12^.

We included participants who met the following criteria: **(i)** residency in Australia at the time of participation; **(ii)** any age; **(iii)** a diagnosis of PD with prescribed medications for PD; and **(iv)** completion of the pain section in the questionnaire. Participants were excluded if they had not completed at least 80% of the questionnaire or did not provide details regarding pain. A full list of questionnaire items included in this analysis is provided in *Supplementary Material 1*. As of the analysis time (October 2024), 10,631 eligible PD participants had completed the questionnaire. Chronic pain was defined as pain persisting for more than three months and occurring on most days or daily.

All participants provided written informed consent, and the information follows the Commonwealth Privacy Act (1988) and National Health and Medical Research Council (NHMRC) Guidelines. The QIMR Berghofer Human Research Ethics Committee granted this study’s ethical approval under project number P3711.

### Study Questionnaire

The baseline questionnaire collected information on **(i)** sociodemographic variables (e.g., age, ethnicity, height, weight, highest qualifications, sex); **(ii)** PD-related variables, including age at PD diagnosis, initial PD symptoms and experiences with memory changes, falls, sleep issues, and other related conditions; **(iii)** pain-related variables including self-reported average chronic pain severity using an 11-point Likert scale (0 = no pain, 10 = most severe pain), identification of specific body sites affected by pain using a body map, and other forms of pain experiences; and **(iv)** presence of 15 medical comorbidities along with lifestyle factors such as smoking, alcohol consumption, and exposure to pesticides or herbicides (see *Supplementary Material, S1*).

### Statistical Analysis

Data analyses were performed using R version 4.3.2 on a MacOS Sonoma 14.4.1 platform. This study investigated the relationship between chronic pain in PD across a range of variables, including sociodemographic and PD-related variables, medical comorbidities, and lifestyle factors. Descriptive statistics were reported as proportions, percentages, frequency distributions, and measures of central tendency. Correlation analyses were conducted using Pearson’s correlation coefficient for continuous variables. Bivariate relationships were assessed using Chi-squared and Fisher’s exact tests for categorical variables, as well as independent samples t-tests for continuous variables. Statistical significance was defined as p < 0.05.

We conducted multiple logistic regression analyses using a backward stepwise approach to identify factors associated with chronic pain. The baseline multivariable model included all variables with a p < 0.20. Multicollinearity among explanatory variables was assessed using the variance inflation factor (VIF). Variables were retained in the model if they had a p < 0.05 and improved the model’s fit, evaluated using the Akaike Information Criterion (AIC). The linearity of continuous variables was evaluated using multivariable fractional polynomial models and their plots. Missing data were addressed by restricting analyses to participants with complete data for all explanatory variables included in the model. Model fit was assessed using the Hosmer-Lemeshow goodness-of-fit test, while classification tables and the area under the receiver operating characteristic (ROC) curve were used to evaluate predictive performance. Results were reported as odds ratios (OR) with 95% confidence intervals (CI) for each risk factor associated with chronic pain.

## RESULTS

### Prevalence and Characteristics of Chronic Pain in PD

Among the 10,631 participants in the cohort, 7,036 (66.2%) reported experiencing chronic pain, with a higher prevalence in females (70.8%, 2,782/3,930) than males (63.5%, 4,254/6,701, p<0.05). At enrollment, the mean age of participants was 70.9 years (± 9.1), with an average age of PD symptom onset at 63.9 years (± 10.6), followed by formal PD diagnosis at 65.6 years (± 9.9) and initiation of levodopa treatment at 66.1 years (± 10.2). Participants with chronic pain were, on average, younger than those without chronic pain (70.3 ± 9.2 vs. 72.3 ± 9.0, p < 0.05), and they experienced symptom onset and began levodopa treatment at younger ages (Table 1). Age patterns were similar across both sexes, regardless of chronic pain status. Participants reporting chronic pain also had a higher weight and BMI compared to those without chronic pain. Also, a significant height difference was noted, with individuals experiencing chronic pain being slightly shorter on average. This trend was consistent across male and female participants (Table 1). Educational attainment was also analysed by gender. Among males, 6,668 out of 6,701 participants provided education data, with 26.7% having completed secondary education and 26.1% having pursued vocational education and training (VET). Similarly, among females, the highest proportions had completed secondary education (31.3%, 1,225/3,914) and VET (24.3%, 950/3,914) (Table 1).

**Table 1.** Demographic characteristics of study participants.

| Variables | All | PD with chronic pain | PD without chronic pain |
| --- | --- | --- | --- |
| Total (n, %) | 10631 | 7036 (66.2) | 3595 (33.8) |
| Age (mean [SD]) | 70.9 ± 9.1 | 70.3 ± 9.2 | 72.3 ± 9 * |
| Age at onset (mean [SD]) | 63.9 ± 10.6 | 62.9 ± 10.8 | 66.1 ± 10.2 * |
| Age of PD diagnosis (mean [SD]) | 65.6 ± 9.9 | 64.1 ± 9.1 | 66.6 ± 9.0* |
| Age of treatment with levodopa (mean [SD]) | 66.1 ± 10.2 | 65.0 ± 10.2 | 68.1 ± 9.7 * |
| BMI (kg/m2) | 27.2 ± 5.4 | 27.6 ± 5.6 | 26.2 ± 4.8 * |

| Variables | All Male | PD with chronic pain | PD without chronic pain |
| --- | --- | --- | --- |
| Total (n, %) | 6701 (63) | 4254 (63.5) | 2447 (36.5) |
| Age (mean [SD]) | 71.2 ± 9.1 | 70.5 ± 9.1 | 72.4 ± 8.9 * |
| Age at onset (mean [SD]) | 64.3 ± 10.6 | 63.2 ± 10.7 | 66.3 ± 10.1 * |
| Age of PD diagnosis (mean [SD]) | 65.6 ± 9.8 | 64.2 ± 9.9 | 67.1 ± 9.1* |
| Age of treatment with levodopa (mean [SD]) | 66.3 ± 10.1 | 65.1 ± 10.2 | 68.2 ± 9.7 * |
| Weight (kg) | 83.0 ± 15.8 | 85.1 ± 16.1 | 81.7 ± 15.0 * |
| Height (cm) | 175.4 ± 8.0 | 175.2 ± 8.2 | 175.6 ± 7.6 |
| BMI (kg/m2) | 27.3 ± 5.0 | 27.8 ± 5.2 | 26.5 ± 4.5 * |
| Education (n, %) | 6668 |  |  |
| Primary Education (1st to 7th grade) | 221 (3.3) | 142 (3.4) | 79 (3.3) |
| Junior Secondary Education (8th and 9th grade) | 543 (8.1) | 372 (8.8) | 171 (7.1) |
| Secondary Education (10th to 12th grade) | 1781 (26.7) | 1174 (27.7) | 607 (25.1) |
| Vocational Education and Training (VET) | 1742 (26.1) | 1144 (27.0) | 589 (24.3) |
| Undergraduate Education | 1021 (15.3) | 605 (14.3) | 416 (17.2) |
| Postgraduate Education | 1360 (20.4) | 799 (18.9) | 561 (23.2) |

| Variables | All Female | PD with chronic pain | PD without chronic pain |
| --- | --- | --- | --- |
| Total | 3930 (37) | 2782 (70.8) | 1148 (29.2) |
| Age (mean [SD]) | 70.5 ± 9.3 # | 69.8 ± 9.2 | 72 ± 9.2 * |
| Age at onset (mean [SD]) | 63.4 ± 10.7 # | 62.5 ± 10.8 | 65.7 ± 10.4 * |
| Age of PD diagnosis (mean [SD]) | 64 ± 10 # | 63.1 ± 10.9 | 65.1 ± 9.8 * |
| Age of treatment with levodopa (mean [SD]) | 65.7 ± 10.2 | 64.9 ± 10.4 | 67.8 ± 9.8 * |
| Weight (kg) | 70.2 ± 16.7 # | 71.8 ± 17.1 | 66.6 ± 15.1 * |
| Height (cm) | 161.6 ± 7.5 # | 161.7 ± 7.5 | 161.3 ± 7.6 |
| BMI (kg/m2) | 26.2 ± 6.0 # | 27.4 ± 6.2 | 25.5 ± 5.5 * |
| Education (n, %) | 3914 |  |  |
| Primary Education (1st to 7th grade) | 100 (2.6) | 65 (2.3) | 35 (3.1) |
| Junior Secondary Education (8th and 9th grade) | 348 (8.9) | 245 (8.9) | 103 (9.0) |
| Secondary Education (10th to 12th grade) | 1225 (31.3) | 857 (31.0) | 368 (32.1) |
| Vocational Education and Training (VET) | 950 (24.3) | 703 (25.4) | 247 (21.6) |
| Undergraduate Education | 529 (13.5) | 371 (13.4) | 158 (13.8) |
| Postgraduate Education | 762 (19.5) | 527 (19.0) | 235 (20.5) |

PD, Parkinson's disease; SD, standard deviation; BMI, body mass index; \* PD with chronic pain vs PD without chronic pain $p < 0.05$ ; # female vs male $p < 0.05$

Chronic pain was the index non-motor symptom in only 1.28% (106/8,305) of cases, with a prevalence of 1.3% (69/5,312) in males and 1.24% (37/2,993) in females, with tremors being the most common initial symptom overall. The average pain severity score among those with chronic pain was 4.4 ± 2.2 out of 10, with females reporting higher pain intensity than males (4.7 ± 2.2 vs. 4.3 ± 2.2, p < 0.05) (Fig 1). Pain was reported bilaterally across 66 specific body regions, with the buttocks (right: 35.6%, left: 32.6%), lower back (right: 24.5%, left: 22.2%), back of the neck (right: 14.1%, left: 14.5%), and front of the knees (right: 14.2%, left: 13.5%) being most frequently affected (Fig 2, Supplementary Material, S2). A *Poisson* regression indicated that higher pain severity score was associated with a greater number of body sites affected (p < 0.05), while being male was associated with slightly fewer affected regions (p = 0.007). Participants reported various pain types, including motor-related, joint, and neuropathic pain (e.g., burning or pins-and-needles), with females more frequently endorsing all categories (Table 2).

**Figure 1.**
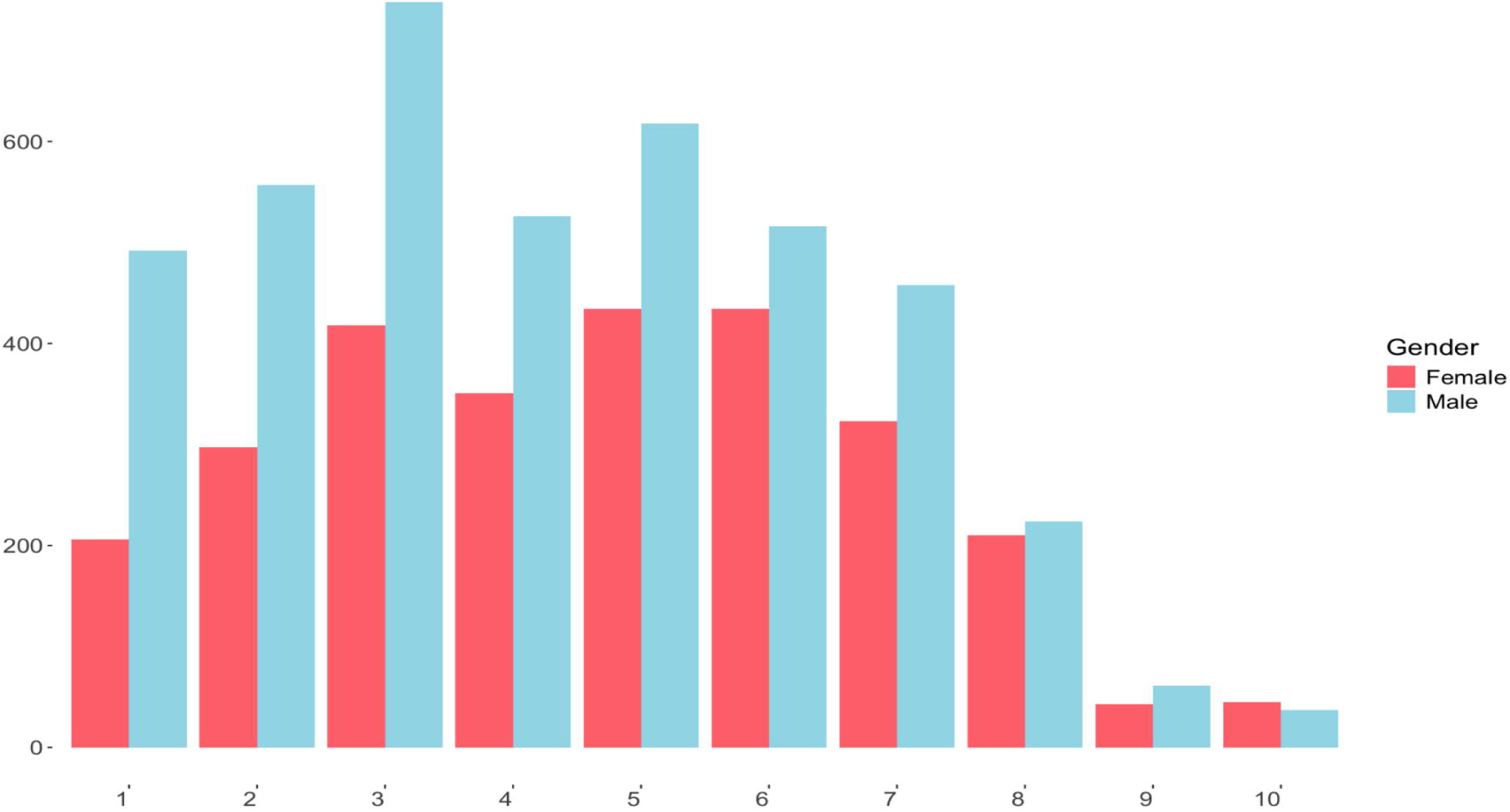
Chronic pain severity. This histogram illustrates the distribution of chronic pain severity scores (0–10) reported by participants in the cohort who experienced persistent pain (n = 7,036). The x-axis denotes pain severity scores, and the y-axis shows the number of participants reporting each value. Males are indicated by blue bars and females by red bars, enabling comparison of pain severity distributions between genders.

**Figure 2.**
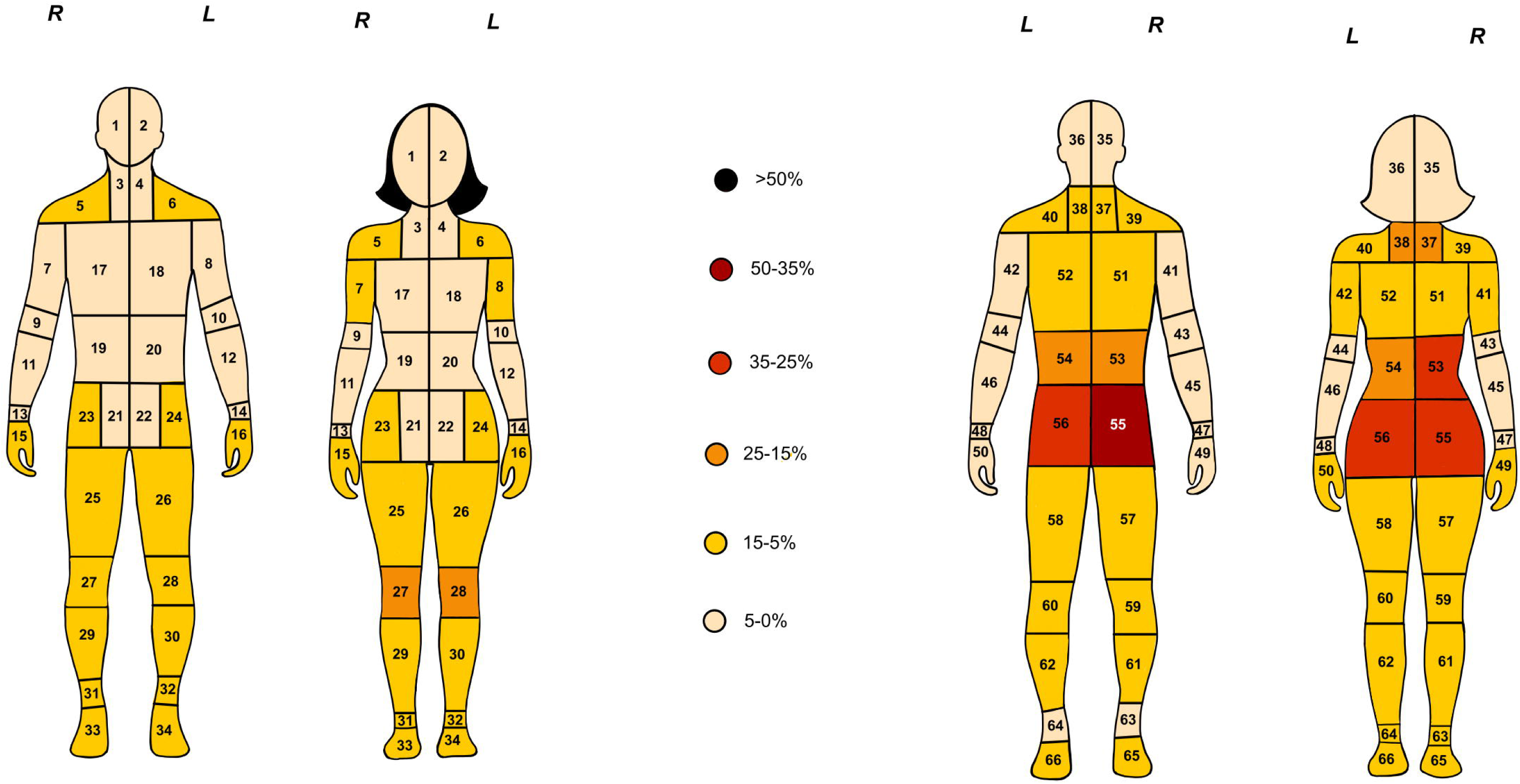
Anatomical distribution of chronic pain. This figure shows the anatomical distribution of chronic pain based on participant self-reports. The body maps, presented in both front and back views, are divided into 66 distinct regions on both the right (R) and left (L) sides. Each region is colour-coded according to the percentage of participants who reported experiencing pain in that area, with the following intervals: 50–35%, 35–25%, 25–15%, 15–5%, and 5–0%.

**Table 2.**
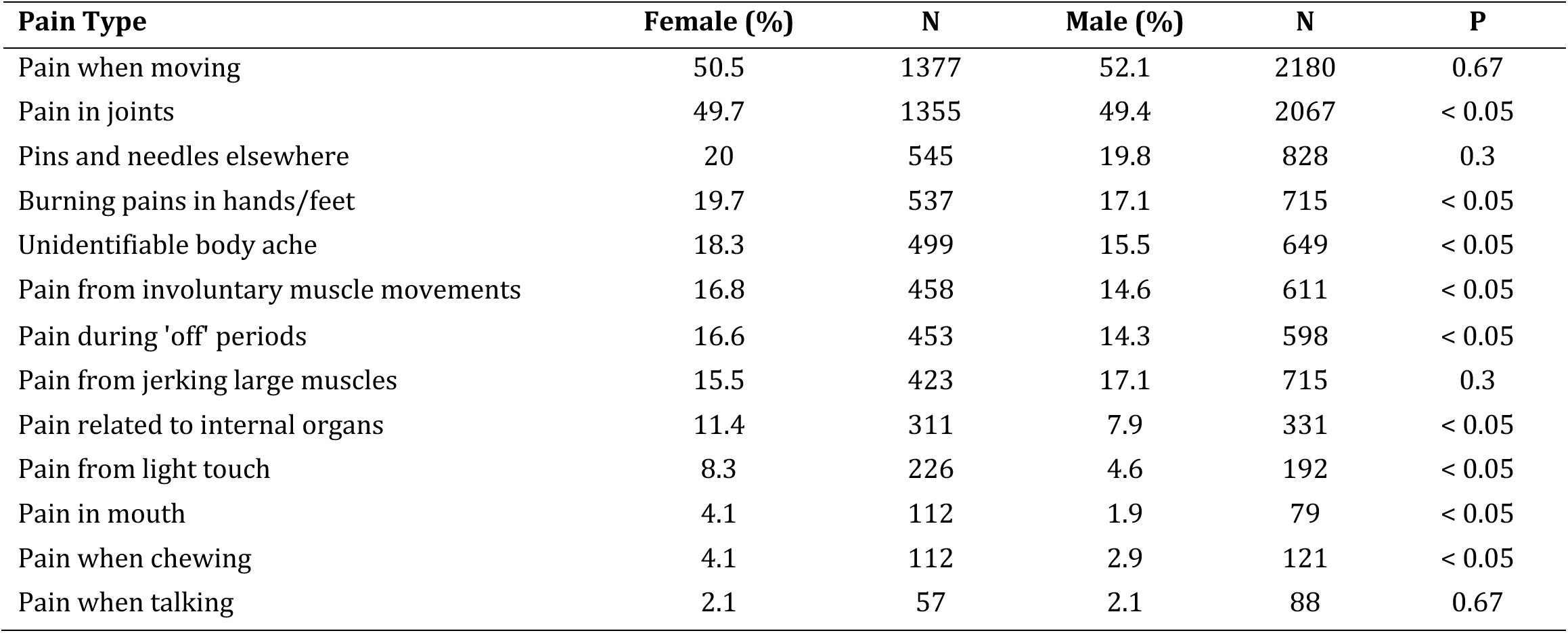
Other Forms of chronic pain reported in the APGS cohort.

| **Pain Type** | **Female (%)** | **N** | **Male (%)** | **N** | **P** |
| --- | --- | --- | --- | --- | --- |
| Pain when moving | 50.5 | 1377 | 52.1 | 2180 | 0.67 |
| Pain in joints | 49.7 | 1355 | 49.4 | 2067 | < 0.05 |
| Pins and needles elsewhere | 20 | 545 | 19.8 | 828 | 0.3 |
| Burning pains in hands/feet | 19.7 | 537 | 17.1 | 715 | < 0.05 |
| Unidentifiable body ache | 18.3 | 499 | 15.5 | 649 | < 0.05 |
| Pain from involuntary muscle movements | 16.8 | 458 | 14.6 | 611 | < 0.05 |
| Pain during 'off' periods | 16.6 | 453 | 14.3 | 598 | < 0.05 |
| Pain from jerking large muscles | 15.5 | 423 | 17.1 | 715 | 0.3 |
| Pain related to internal organs | 11.4 | 311 | 7.9 | 331 | < 0.05 |
| Pain from light touch | 8.3 | 226 | 4.6 | 192 | < 0.05 |
| Pain in mouth | 4.1 | 112 | 1.9 | 79 | < 0.05 |
| Pain when chewing | 4.1 | 112 | 2.9 | 121 | < 0.05 |
| Pain when talking | 2.1 | 57 | 2.1 | 88 | 0.67 |

Univariate analyses also revealed higher prevalence of chronic pain among certain comorbid conditions, including those who report having sleep apnoea, depression, osteoarthritis, mild cognitive impairment, osteoporosis, inflammatory bowel disease, rheumatoid arthritis, essential tremor, hypothyroidism, high blood pressure, REM sleep behaviour disorder, and restless legs syndrome (p<0.05). On the other hand, we identified significant associations between chronic pain and lifestyle exposures such as exposure to pesticides, herbicides, and heavy metals, consumption of alcohol and caffeinated soft drinks (p<0.05) (Tables 3–4). However, these associations may reflect confounding or shared risk factors.

**Table 3.** Chronic pain concerning medical comorbidities in the APGS cohort.

| Clinical variables | PD with chronic pain |  | PD without chronic pain |  | P |
| --- | --- | --- | --- | --- | --- |
|  | N | % | N | % |  |
| Depression | 2337 | 33.2 | 705 | 19.6 | < 0.05 |
| Sleep Apnoea | 1201 | 17.1 | 448 | 12.56 | < 0.05 |
| Restless Leg Syndrome | 1109 | 15.8 | 493 | 13.7 | < 0.05 |
| Essential Tremor | 879 | 12.5 | 388 | 10.8 | 0.05 |
| High Blood Pressure | 875 | 12.4 | 54 | 1.5 | < 0.05 |
| REM Sleep Behaviour Disorder | 833 | 11.8 | 270 | 7.5 | < 0.05 |
| Osteoarthritis | 680 | 9.7 | 26 | 0.7 | < 0.05 |
| Mild Cognitive Impairment | 148 | 2.1 | 14 | 0.4 | < 0.05 |
| Osteoporosis | 248 | 3.5 | 15 | 0.4 | < 0.05 |
| Psoriasis | 141 | 2 | 8 | 0.2 | 0.59 |
| Hypothyroidism | 130 | 1.8 | 6 | 0.2 | < 0.05 |
| Inflammatory Bowel Disease (IBD) | 110 | 1.6 | 8 | 0.2 | < 0.05 |
| Peptic Ulcer | 127 | 1.8 | 5 | 0.1 | 0.27 |
| Rheumatoid Arthritis | 111 | 1.6 | 7 | 0.2 | < 0.05 |
| Stroke | 112 | 1.6 | 10 | 0.3 | 0.77 |

**Table 4.** Environmental and lifestyle risk factors in the APGS cohort.

| Lifestyle Factors | PD with chronic pain |  | PD without chronic pain |  | P |
| --- | --- | --- | --- | --- | --- |
|  | N | % | N | % |  |
| Exposed to pesticides/herbicides | 2716 | 38.6 | 1194 | 33.2 | < 0.05 |
| Heavy metal poisoning | 86 | 1.2 | 23 | 0.6 | < 0.05 |
| Drinks caffeinated coffee | 5335 | 75.8 | 2714 | 75.5 | 0.72 |
| Drinks tea | 4906 | 69.7 | 2523 | 70.2 | 0.65 |
| Drinks alcohol | 4775 | 22.9 | 2522 | 18.4 | < 0.05 |
| Ever been a regular smoker (min. 12 months) | 179 | 67.9 | 53 | 70.2 | < 0.05 |
| Drinks caffeinated soft drinks | 1612 | 2.5 | 662 | 1.5 | < 0.05 |

| Lifestyle Factors | Female PD with chronic pain |  | Male PD with chronic pain |  | P |
| --- | --- | --- | --- | --- | --- |
|  | N | % | N | % |  |
| Exposed to pesticides/herbicides | 826 | 29.7 | 1890 | 44.4 | < 0.05 |
| Heavy metal poisoning | 31 | 1.1 | 55 | 1.3 | 0.58 |
| Drinks caffeinated coffee | 1994 | 71.7 | 3341 | 78.5 | < 0.05 |
| Drinks tea | 2048 | 73.6 | 2858 | 67.2 | < 0.05 |
| Drinks alcohol | 1627 | 58.5 | 3148 | 74 | < 0.05 |
| Ever been a regular smoker (min. 12 months) | 62 | 2.2 | 117 | 2.8 | 0.2 |
| Drinks caffeinated soft drinks | 495 | 17.8 | 1117 | 26.3 | < 0.05 |

### Multivariable Associations of Chronic Pain

Multivariable logistic regression identified several independent predictors of chronic pain after adjusting for potential confounders, as shown in Table 5. Twelve candidate variables with a p<0.20 in univariate analysis were included in the initial multivariable model, followed by stepwise backward elimination, yielding a final model with eight retained variables.

**Table 5.**
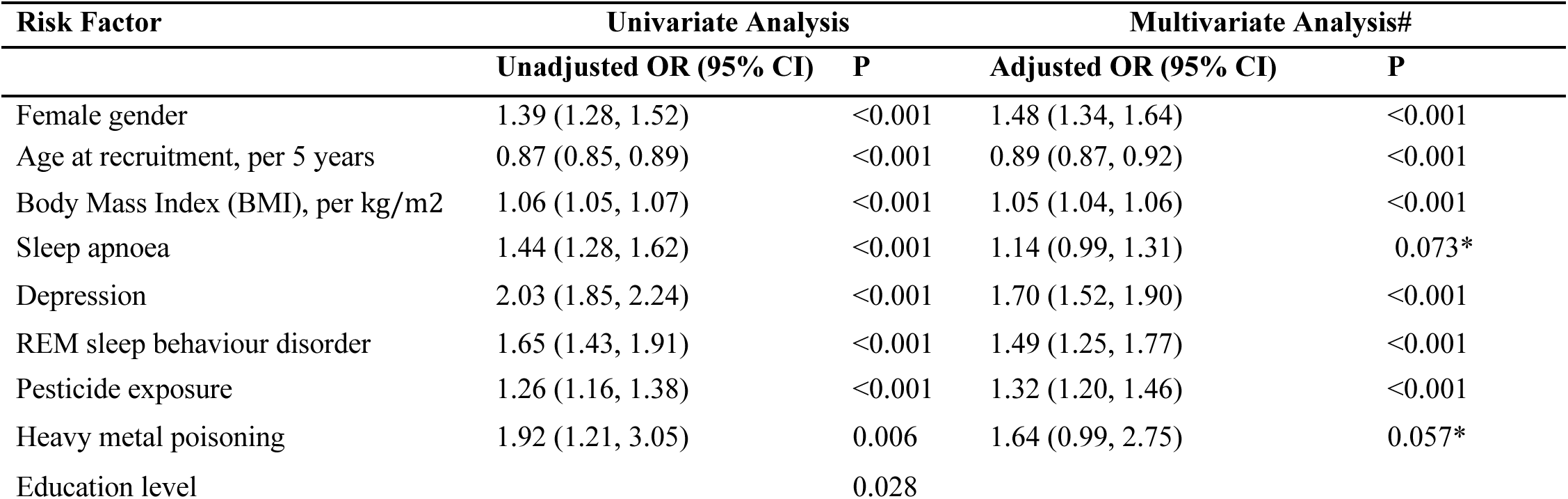
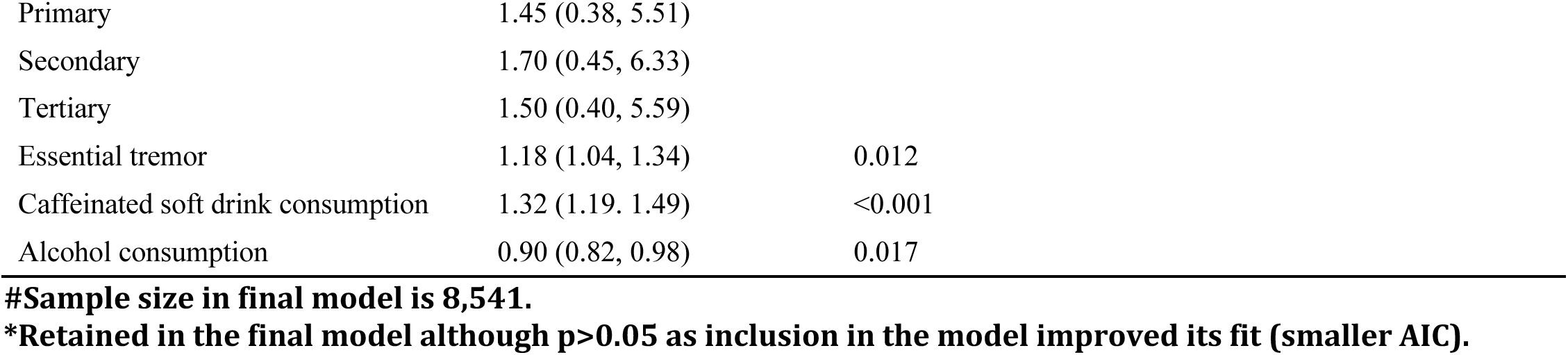
Univariate and multivariate logistic regression analyses.

| Risk Factor | Univariate Analysis |  | Multivariate Analysis# |  |
| --- | --- | --- | --- | --- |
|  | Unadjusted OR (95% CI) | P | Adjusted OR (95% CI) | P |
| Female gender | 1.39 (1.28, 1.52) | <0.001 | 1.48 (1.34, 1.64) | <0.001 |
| Age at recruitment, per 5 years | 0.87 (0.85, 0.89) | <0.001 | 0.89 (0.87, 0.92) | <0.001 |
| Body Mass Index (BMI), per kg/m<sup>2</sup> | 1.06 (1.05, 1.07) | <0.001 | 1.05 (1.04, 1.06) | <0.001 |
| Sleep apnoea | 1.44 (1.28, 1.62) | <0.001 | 1.14 (0.99, 1.31) | 0.073* |
| Depression | 2.03 (1.85, 2.24) | <0.001 | 1.70 (1.52, 1.90) | <0.001 |
| REM sleep behaviour disorder | 1.65 (1.43, 1.91) | <0.001 | 1.49 (1.25, 1.77) | <0.001 |
| Pesticide exposure | 1.26 (1.16, 1.38) | <0.001 | 1.32 (1.20, 1.46) | <0.001 |
| Heavy metal poisoning | 1.92 (1.21, 3.05) | 0.006 | 1.64 (0.99, 2.75) | 0.057* |
| Education level |  | 0.028 |  |  |

|  |  |  |
| --- | --- | --- |
| Primary | 1.45 (0.38, 5.51) |  |
| Secondary | 1.70 (0.45, 6.33) |  |
| Tertiary | 1.50 (0.40, 5.59) |  |
| Essential tremor | 1.18 (1.04, 1.34) | 0.012 |
| Caffeinated soft drink consumption | 1.32 (1.19, 1.49) | <0.001 |
| Alcohol consumption | 0.90 (0.82, 0.98) | 0.017 |

**#Sample size in final model is 8,541.**
**\*Retained in the final model although $p > 0.05$ as inclusion in the model improved its fit (smaller AIC).**

Female sex, higher BMI, depression, REM sleep behaviour disorder, and prior pesticide exposure were significantly associated with increased odds of chronic pain, highlighting the multifactorial nature of pain susceptibility in PD. Age at recruitment was inversely associated with chronic pain risk, with each five-year increase associated with lower odds of chronic pain even after adjustment. While sleep apnoea and heavy metal poisoning were strongly associated with pain in univariate analyses, their associations attenuated in the multivariable model. However, both were retained due to their contribution to improved model fit, as indicated by lower AIC values.

In contrast, variables such as essential tremor, alcohol consumption, and caffeinated soft drink intake, though significant in univariate models, were not retained after adjustment, suggesting their associations were confounded or mediated by other variables in the model. Education level did not display a consistent gradient across categories and was excluded from the final model, though its initial association may reflect underlying socio-demographic influences on pain perception.

The final model demonstrated modest discriminatory performance, with a C-statistic (AUC) of 0.63 (95% CI 0.62-0.64), and acceptable calibration, as indicated by a non-significant Hosmer-Lemeshow test (p = 0.25). Interaction terms were tested to assess effect modification by sex, but none reached statistical significance; and sex-stratified analyses were therefore not pursued.

## DISCUSSION

The study represents the largest investigation of chronic pain in people living with PD, providing comprehensive insights into its prevalence, severity, and anatomical distribution, as well as its relationship with sociodemographic variables, medical comorbidities, and lifestyle factors. In our cohort, two-thirds of participants reported chronic pain, yet only 1.28% identified it as an initial symptom of PD. Pain was more prevalent and severe in females and frequently involved the buttocks, lower back, neck, and knees. In addition, chronic pain was found to be associated with several medical comorbidities, such as depression, sleep apnoea and restless legs syndrome. Chronic pain was also correlated with environmental and lifestyle factors, such as smoking, alcohol and caffeine intake, and exposure to heavy metals and pesticide exposure, with a significantly higher exposure rate in males.

The prevalence of chronic pain was 66.2%, within the broad range of 30–85% reported. This variation highlights the heterogeneous nature influenced by geographical and cultural contexts. For instance, high rates (70%-90%) were reported in cohorts from Russia^13^, Norway^14^, Mexico^15^, Japan^16^, the UK^17^ and Italy^18^. Moderate prevalence rates (50%-60%) were found in Turkey^19^, Germany^20^, India^10^ and Brazil^9^. These discrepancies may stem from differences in study design, chronic pain definitions, sample sizes, and population characteristics. To provide context, Miller et al.^21^ and the nationwide report *The Cost of Pain in Australia*^22^ estimated chronic pain prevalence in individuals aged 70 years as 23% in males and 30% in females, making our cohort’s prevalence notably higher than that in the general population. Recent longitudinal studies, such as the one by Gunzler et al. have further demonstrated that individuals with PD exhibit distinct subgroups with shared trajectories of pain severity and persistence over time, influenced by disease duration and symptom burden^23^. While our cross-sectional design does not allow us to assess longitudinal pain patterns, our findings complement this work by highlighting demographic, anatomical, and environmental risk profiles within a large national cohort.

Chronic pain as an initial symptom has been reported in other cohorts with variation across gender, location, origin, and age^24^. Gatto et al. linked the *LRRK* G2019S variant to early pain in an Argentinian cohort^25^, while Guiffrida et al. found that 25% of participants reported pain in the early disease stages of PD^26^–a rate much higher than our finding of 1.28%. Consistent with previous studies, females in our cohort reported higher pain severity than males, though the clinically significance of this difference remains uncertain. Genetic factors might contribute to these differences; for instance, the *DRD2* rs2283265 polymorphism has been associated with an increased risk of PD-related pain in females^27^, while the *CHRNA4* rs1044397 variant may influence PD onset age in females with chronic pain^28^.

Our multivariable analysis confirmed that gender is a strong determinant of chronic pain, with females nearly 50% more likely to experience it than males. This disparity may be attributed to differences in sensory processing pain and pain relief responses, influenced by sex chromosomes and hormonal factors such as testosterone and oestradiol, suggesting a genetic and physiological predisposition for PD-related pain in females. Interestingly, younger age was associated with a higher likelihood of reporting chronic pain, which contrasts with studies suggesting increased pain prevalence in older adults. A possible explanation could be that older individuals may develop pain tolerance or underreport pain due to psychosocial factors, such as age-related stoicism or the normalisation of pain as part of aging. In contrast, younger individuals may experience more intense pain due to factors like motor dysfunction, neuroplasticity, and increased psychosocial stress^29^.

On the other hand, our analysis revealed that higher BMI, depression, REM sleep behaviour disorder, and pesticide exposure were independently associated with chronic pain in PD. These findings underscore the multifactorial nature of pain susceptibility and the need for personalised management strategies. In contrast, several variables that showed significant univariate associations, such as alcohol use, caffeine intake, and heavy metal exposure, did not remain significant after adjustment, suggesting their effects may be confounded or mediated by other factors.

Previous studies indicate that pain localisation in PD varies by gender and different ancestries, making it difficult to draw an accurate pain topography. In our cohort, the most commonly reported pain sites were the buttocks (∼35–31% of cases), lower back (∼25–22%), front of the knee (∼17–13%), and back of the neck (∼19–14%), with females reporting more pain. According to the Australian Institute of Health & Welfare (AIHW) 2023 report, the prevalence of back pain in Australia is 16%^24^. While structural changes in the lumbar spine may contribute to back pain^25^, psychosocial factors, including stress and mental health, are stronger predictors^26^. In PD patients, chronic musculoskeletal pain (as defined by the IASP ICD-11 classification) is often exacerbated by dystonia, rigidity, and postural instability^27^, and PD has been associated with decreased pain thresholds, reduced pain tolerance, and inhibition of the substantia gelatinosa’s pain suppression pathways^28,29^. These mechanisms align with nociplastic pain, characterised by altered central pain processing in the absence of tissue damage or structural pathology^30^.

Truncal instability in PD may contribute to chronic back and neck pain^31^. Participants in that study reported chronic pain in an average of four to five anatomical regions, indicating widespread and diverse experiences of pain. Higher pain severity scores were also associated with an increase in the number of body sites affected. Although the current study’s questionnaire did not specifically interrogate the underlying mechanisms of pain (e.g., neuropathic/nociplastic/nociceptive pathways), our results provide valuable clues. For instance, joint-related pain may indicate nociceptive/musculoskeletal mechanisms; burning or pins-and-needles sensations could suggest neuropathic pain^32^; and widespread pain associated with depression or hyperalgesia is consistent with nocicplastic pain^30^. Gender differences in chronic pain may reflect biological and psychosocial factors, with females showing heightened sensitivity due to hormonal fluctuations, opioid system variation, and stress^17^, underscoring the need for personalised, multidisciplinary pain management in PD. Furthermore, although the current study did not include formal clinical assessments of motor or non-motor fluctuations, approximately 17% of participants reported pain during “off” periods or in association with involuntary movements and large muscle jerks, symptoms suggestive of underlying motor fluctuations. Pain related to fluctuating motor states, particularly “off-period” pain, is a recognised subtype of PD-related pain and is often responsive to dopaminergic therapy. While the cross-sectional design and questionnaire-based methodology limited our ability to characterise these phenomena in detail, the presence of these specific pain descriptors highlights the clinical relevance of symptom fluctuations in shaping the pain experience in PD.

The prevalence, severity, and types of chronic pain vary with disease stage and medical comorbidities. The comorbidity burden increases as PD progresses, underscoring the necessity of ongoing clinical assessment and management^33^. Our study identified significant associations between chronic pain and multiple comorbidities, though causal relationships remain uncertain. For instance, 33.2% of PD participants with chronic pain reported depression, suggesting a bidirectional relationship^34^. Depression can alter the perception of PD-related pain and accelerate its progression to a severe and refractory state^35^. Recent evidence suggests that glutamatergic dysregulation may serve as a common neurobiological mechanism linking depression and chronic pain, particularly through abnormal excitatory neurotransmission and central sensitisation^36^ (Pagonabarraga et al., 2021). This shared pathophysiology may amplify both affective and sensory components of the pain experience in PD, warranting further investigation into glutamate-targeting therapies.

Sleep disorders, including REM sleep behaviour disorder and sleep apnoea, were also common, with chronic pain negatively impacting sleep quality and leading to sleep fragmentation and insomnia^37^. Musculoskeletal conditions such as osteoarthritis and osteoporosis, reported by 4% of participants, contribute to the pain experienced by 40%–75% of PD participants, often linked to truncal deformities and bone mineralisation disorders^38^.

Furthermore, 10% of participants in the current study reported comorbid osteoarthritis—a condition linked to specific pain characteristics, including paresthesia and akathisia-related pain^39^. Other comorbidities observed included rheumatoid arthritis and inflammatory bowel disease, which may exacerbate PD-related pain through inflammatory mechanisms. Restless leg syndrome, essential tremor, and mild cognitive impairment, associated with dopamine deficiency and disrupted brain circuitry, can further exacerbate pain in PD. For example, changes in nociceptive processing and regulation have been observed in early PD, which may be related to the same dopaminergic dysfunction that causes tremors. The basal ganglia, involved in the generation of Parkinsonian tremors, also play a role in pain processing, potentially intensifying symptoms in affected individuals^40^.

PD participants reporting chronic pain in our cohort exhibited higher rates of alcohol consumption, coffee and tea drinking, smoking, and exposure to pesticides, herbicides, and heavy metals, consistent with existing literature^41^. Smoking and alcohol use have been linked to chronic pain, with smoking-associated vascular damage and systemic inflammation potentially contributing to increased pain risk^42^. Alcohol exerts neurotoxic effects on both central and peripheral nerves, leading to harmful changes in neural structures and musculoskeletal systems, while excessive consumption is associated with various chronic pain conditions and poorer pain outcomes. The relationship between substance use and chronic pain is complex and potentially bidirectional, as individuals may use these substances as coping mechanisms to alleviate pain and discomfort^43^. While coffee and tea consumption have been associated with a reduced risk of developing PD, their relationship with chronic pain is more complex and less well established. Caffeine may exert mild analgesic effects by antagonising adenosine receptors involved in pain modulation; however, excessive intake has been linked to increased anxiety, sleep disturbances, and withdrawal headaches, which may indirectly exacerbate pain^41^. In our study, higher rates of coffee and tea consumption were observed among participants with chronic pain, though this association did not remain significant in multivariable models. This may reflect confounding by other lifestyle or neuropsychiatric factors. On the other hand, pesticide and herbicide exposure, and heavy metal poisoning are known PD risk factors due to their neurotoxic properties, including oxidative stress, neuroinflammation, mitochondrial dysfunction, and dopaminergic neuronal cell death^44^. These same mechanisms are also implicated in the pathophysiology of chronic pain, suggesting a shared biological pathway. Moreover, cumulative exposure to these environmental toxins may exacerbate neurodegeneration and sensitize pain pathways, particularly in individuals with an existing vulnerability such as PD^44^.

Our study has several limitations. First, the recruitment method may have introduced self-selection bias, as individuals experiencing more severe or bothersome symptoms, such as chronic pain, may have been more motivated to participate. This could lead to an overestimation of pain prevalence and severity in our cohort. Additionally, the use of patient-reported questionnaires is subject to recall bias, which may have influenced the accuracy of reported symptom onset, pain characteristics, and lifestyle exposures. Participants with advanced PD, including those with cognitive impairment and severe musculoskeletal disabilities, were less likely to participate. Second, the absence of a control group without PD limits direct comparisons. However, previous studies have shown that pain is prevalent in the general population, allowing for indirect comparisons.^45^ Third, the eligibility criterion requiring antiparkinsonian medication use may have excluded non-medicated individuals, limiting generalisability of our findings to the pain experiences of non-medicated PD individuals. Fourth, although our questionnaire covered a broad range of symptoms, it did not employ a validated PD-specific pain scale. This limits the standardisation and comparability of our pain assessments with other studies and may have impacted the classification of pain subtypes. Fifth, the cross-sectional design of the study precludes conclusions about causality. Some associations observed, for example with depression or lifestyle factors, may reflect reverse causation, whereby chronic pain influences those outcomes rather than results from them. Sixth, we did not explicitly collect data on menopausal status. Given that most female participants were aged over 60, it is likely that the majority were postmenopausal. However, the lack of this information limits our ability to assess the influence of hormonal transitions, such as menopause, on pain sensitivity and perception in PD. Hormonal status, particularly the role of oestrogens, may play a key role in shaping pain experiences in females. Lastly, the lack of detailed information on pain types and pain treatments limits the accurate assessment of chronic pain characteristics in our cohort. The effects of antiparkinsonian medications on pain remain to be reported.

In conclusion, chronic pain is a complex and highly prevalent feature of PD, affecting approximately two-thirds of individuals and often co-occurring with comorbidities and environmental risk factors. Younger participants and females reported more frequent and severe pain, while anatomical distribution patterns and diverse pain types highlight the complexity of pain phenotypes in PD. These findings emphasise the urgent need for improved clinical recognition, comprehensive assessment, and tailored management strategies that integrate medical, psychosocial, and lifestyle interventions. Importantly, they also stress the value of addressing modifiable risk factors, such as mental health conditions and environmental exposures, in shaping targeted prevention and therapeutic approaches. Future research should focus on elucidating the underlying mechanisms driving chronic pain variability in PD and developing evidence-based guidelines to enhance patient quality of life and treatment outcomes.

## Acknowledgments

We are deeply grateful to all the participants of the Australian Parkinson’s Genetics Study (APGS) whose invaluable contributions and commitment made this research possible. APGS is supported by the Shake It Up Australia Foundation and the Michael J. Fox Foundation for Parkinson’s Research (MJFF-021952). MER thanks the support from the Rebecca L. Cooper Medical Research Foundation (F20231230). LMGM is supported by a UQ Research Training Scholarship from the University of Queensland (UQ). KRK is supported by grants from the Medical Research Future Fund and the Ainsworth 4 Foundation (unrelated to the current study). JA is supported by grants from the National Health & Medical Research Foundation (unrelated to the current study).

## Author Contributions

N.S.O., F.Ch., and M.E.R. conceived the study. N.S.O. and F.Cao. curated the data. N.S.O. conducted the statistical analyses under the direction and supervision of M.E.R. N.S.O. and F.Ch. prepared the initial draft and incorporated feedback from all co-authors. M.E.R. designed the cohort study, oversaw participant recruitment and data collection for APGS, and obtained funding for the study. T.T.N. designed the pain module for genetic and epidemiological investigations as part of a broader Precision Pain Medicine R&D program. All authors contributed to data interpretation, critically reviewed each draft of the report, and approved the final submitted version.

## Data availability

The data supporting the findings of this study are not publicly accessible due to participant privacy considerations. However, anonymised datasets can be made available to qualified researchers upon request to the corresponding author. Data sharing will be contingent upon obtaining all necessary ethical approvals and signing an institutional data transfer agreement.

