## Supplementary Material for "Chronic Pain in Parkinson’s Disease: Prevalence, Sex Differences, Regional Anatomy and Comorbidities"

This file contains the following supplementary information for the manuscript:

1. Graphical Abstract
2. Questions included in this epidemiological study
3. Anatomical distribution of chronic pain in the APGS cohort

**S1. Graphical Abstract**

Chronic pain affects 66.2% of individuals with Parkinson’s disease (PD), with a higher prevalence in females (70.8%). Chronic pain is most commonly reported in the buttocks (35.6%), lower back (25.4%), neck (19.4%), and knees (17.2%). Severity is associated with several comorbidities, including depression, sleep disorders, and osteoarthritis, among others. Environmental exposures, including pesticides and heavy metals, increase pain prevalence, particularly in males. These findings highlight sex differences and multimorbidity associated with PD.

**S2. Questions included in this epidemiological study**

a. **Sociodemographic questions:**

- Are you male or female?
- How old are you?
- What is your ethnicity?
- What is your ancestry?
- What is your current height (cm)?
- What is your current weight? (kg)?
- What is your highest qualification?

b. **PD related questions:**

- Do you have a current diagnosis of Parkinson’s disease?
- How old were you approximately when you or those close to you first noticed the onset of Parkinson’s symptoms?
- How old were you when you were formally diagnosed with Parkinson's disease by a healthcare professional?
- What was your first symptom of Parkinson's disease?
- Have you experienced a reduction or loss of the sense of smell related to Parkinson's disease?
- Have you ever experienced falls as a result of Parkinson’s disease?
- Have you ever experienced light-headedness or dizziness when standing up from sitting or in lying position?
- Have you experienced changes to your memory?
- Have you ever experienced sleep problems?
- How old were you when you started taking levodopa?

c. **Pain-related questions:**

- On the diagram, please click or tap on any other areas where you experience pain: head, neck, shoulder, upper arm, elbow, forearm, lower arm, wrist, hand, chest, abdomen, upper back, lower back, groin, bottom, hip, upper leg, knee, lower leg, ankle, foot (right and left, front and back).
- Do you have chronic pain or pain lasting for more than 3 months?
- Do you have pain right now?
- On a scale of 0 - 10, what is your pain on average?
- How would you rate your pain right now?
- Do you have pain in your mouth?
- Do you experience pain when chewing?
- Do you have pain when talking?
- Do you have pain in your joints?
- Do you have pain when trying to move?
- Do you have pain related to internal organs like the stomach, liver, kidneys, or bowels?
- Do you have an ache in your body that you cannot describe?
- Do you feel burning pains in your hands and/or feet?
- Do you have pins and needles anywhere else on your body?
- Do you experience pain related to light touch?
- Do you have pain related to involuntary small muscle movements?
- Do you have pain related to jerking big muscle movements?
- Do you experience pain during the “off” periods when you cannot move?

d. **Medical Comorbidities questions:**

- Have you ever had REM Sleep Behaviour Disorder?
- Have you ever had Restless legs syndrome?
- Have you ever had Sleep apnea?
- Have you ever had Depression?
- Have you ever had High Blood Pressure?
- Have you ever had Osteoarthritis?
- Have you ever had Psoriasis?
- Have you ever had Hypothyroidism?
- Have you ever had Essential Tremor?
- Have you ever had Mild cognitive impairment?
- Have you ever had Rheumatoid Arthritis?
- Have you ever had Stroke?
- Have you ever had Peptic Ulcer?
- Have you ever had Inflammatory Bowel Disease?
- Have you ever had Osteoporosis?

e. **Lifestyle factors questions:**

- Have you ever been exposed to pesticides or herbicides?
- Have you ever had heavy metal poisoning?
- Do you drink caffeinated coffee (not decaf)?
- Do you drink tea (not decaffeinated)?
- Do you drink caffeinated soft drink (e.g., Coke, Redbull)?
- Do you drink alcohol (e.g. beer, wine, spirits)?
- Do you, or have you ever, smoked cigarettes, cigars, or pipes, at least once a day for one year?

**S3. Anatomical distribution of chronic pain in the APGS cohort.** This table presents the anatomical distribution of chronic pain reported by participants in the Australian Parkinson’s Genetics Study (APGS) cohort. Pain locations are categorised across 66 specific body regions, separately for males and females, and divided into left/right and front/back when applicable. For each region, the absolute number of individuals reporting pain (N) and the corresponding percentage (%) within each sex group are provided. Percentages reflect the proportion of participants within each sex who reported pain in that specific body site.

| **Anatomical** | | | **Reference** | **Female** | | **Male** | |
| --- | --- | --- | --- | --- | --- | --- | --- |
|  |  |  |  | **N** | **%** | **N** | **%** |
| Head | Right | Front | 1 | 3.5 | 96 | 2.3 | 99 |
| Head | Left | Front | 2 | 3.7 | 103 | 2.1 | 90 |
| Neck | Right | Front | 3 | 4.8 | 134 | 3.5 | 150 |
| Neck | Left | Front | 4 | 4.6 | 127 | 3.7 | 158 |
| Shoulder | Right | Front | 5 | 12.6 | 351 | 9.3 | 397 |
| Shoulder | Left | Front | 6 | 11.5 | 321 | 8.6 | 367 |
| Upper Arm | Right | Front | 7 | 5.9 | 165 | 4.7 | 198 |
| Upper Arm | Left | Front | 8 | 6.1 | 169 | 4.8 | 206 |
| Elbow | Right | Front | 9 | 2.9 | 82 | 3 | 128 |
| Elbow | Left | Front | 10 | 3.2 | 90 | 2.4 | 101 |
| Forearm | Right | Front | 11 | 3.6 | 99 | 2.9 | 122 |
| Forearm | Left | Front | 12 | 3.7 | 103 | 2.8 | 119 |
| Wrist | Right | Front | 13 | 3.6 | 99 | 2.5 | 106 |
| Wrist | Left | Front | 14 | 3.4 | 95 | 2.3 | 99 |
| Hand | Right | Front | 15 | 8.2 | 228 | 5.8 | 246 |
| Hand | Left | Front | 16 | 8.3 | 231 | 5.4 | 231 |
| Chest | Right |  | 17 | 1.9 | 53 | 2.1 | 88 |
| Chest | Left |  | 18 | 1.7 | 46 | 1.8 | 75 |
| Abdomen | Right |  | 19 | 3.3 | 93 | 2.3 | 96 |
| Abdomen | Left |  | 20 | 3.5 | 96 | 2.1 | 91 |
| Groin | Right |  | 21 | 3.3 | 91 | 4 | 172 |
| Groin | Left |  | 22 | 3.1 | 87 | 3.6 | 154 |
| Hip | Right |  | 23 | 9.1 | 253 | 6.9 | 294 |
| Hip | Left |  | 24 | 9.4 | 262 | 6.8 | 289 |
| Upper Leg | Right | Front | 25 | 10 | 279 | 7.9 | 334 |
| Upper Leg | Left | Front | 26 | 9.2 | 256 | 8 | 342 |
| Knee | Right | Front | 27 | 17.2 | 479 | 14.2 | 606 |
| Knee | Left | Front | 28 | 16.1 | 449 | 13.5 | 573 |
| Lower Leg | Right | Front | 29 | 7.7 | 213 | 7.3 | 310 |
| Lower Leg | Left | Front | 30 | 8.1 | 225 | 7.3 | 309 |
| Ankle | Right | Front | 31 | 6.6 | 183 | 5 | 211 |
| Ankle | Left | Front | 32 | 6 | 166 | 5.2 | 221 |
| Foot | Right | Front | 33 | 11.5 | 319 | 8.9 | 378 |
| Foot | Left | Front | 34 | 12.2 | 339 | 9.4 | 401 |
| Head | Right | Back | 35 | 4.3 | 119 | 2.4 | 102 |
| Head | Left | Back | 36 | 3.8 | 105 | 2.3 | 96 |
| Neck | Right | Back | 37 | 19.4 | 541 | 14.1 | 598 |
| Neck | Left | Back | 38 | 19 | 528 | 14.5 | 617 |
| Shoulder | Right | Back | 39 | 14.6 | 407 | 9.3 | 396 |
| Shoulder | Left | Back | 40 | 12.8 | 356 | 8.1 | 344 |
| Upper Arm | Right | Back | 41 | 6.1 | 171 | 3.9 | 167 |
| Upper Arm | Left | Back | 42 | 5.4 | 149 | 3.5 | 148 |
| Elbow | Right | Back | 43 | 2.5 | 69 | 2.3 | 96 |
| Elbow | Left | Back | 44 | 2.8 | 77 | 2 | 86 |
| Lower Arm | Right | Back | 45 | 3 | 84 | 2 | 87 |
| Lower Arm | Left | Back | 46 | 2.9 | 80 | 1.8 | 78 |
| Wrist | Right | Back | 47 | 2.2 | 61 | 1.6 | 67 |
| Wrist | Left | Back | 48 | 2.4 | 68 | 1.4 | 58 |
| Hand | Right | Back | 49 | 6.4 | 177 | 4.4 | 186 |
| Hand | Left | Back | 50 | 6.5 | 181 | 4.1 | 176 |
| Upper | Right | Back | 51 | 14.5 | 403 | 9.4 | 399 |
| Upper | Left | Back | 52 | 12.7 | 353 | 8.9 | 377 |
| Lower | Right | Back | 53 | 25.4 | 708 | 24.5 | 1044 |
| Lower | Left | Back | 54 | 22.6 | 628 | 22.2 | 944 |
| Bottom | Right |  | 55 | 34 | 946 | 35.6 | 1516 |
| Bottom | Left |  | 56 | 31.8 | 886 | 32.6 | 1385 |
| Upper Leg | Right | Back | 57 | 8.6 | 238 | 6.9 | 295 |
| Upper Leg | Left | Back | 58 | 8 | 223 | 6.9 | 293 |
| Knee | Right | Back | 59 | 8.6 | 239 | 7 | 296 |
| Knee | Left | Back | 60 | 8 | 222 | 6.8 | 290 |
| Lower Leg | Right | Back | 61 | 9 | 249 | 7.9 | 337 |
| Lower Leg | Left | Back | 62 | 9 | 250 | 8.1 | 343 |
| Ankle | Right | Back | 63 | 5.9 | 165 | 4.1 | 175 |
| Ankle | Left | Back | 64 | 5.9 | 164 | 4.3 | 182 |
| Foot | Right | Back | 65 | 6.2 | 172 | 5.1 | 219 |
| Foot | Left | Back | 66 | 6.6 | 184 | 5 | 223 |
